# Universal Rule for Covid 19 and Herd Immunity in the US

**DOI:** 10.1101/2021.03.05.21251577

**Authors:** Jacob G. Kuriyan

**Affiliations:** Care Maps Inc. and Physmark Inc.

## Abstract

A new Universal rule for Covid 19 data is derived in this paper using the SIR model.

It relates infection and removal rates and is validated by the global Covid 19 data. Over 186,000 data points, from 190 countries and the states of the US, for the period April 1 to December 12, 2020 - fall on a single line, as the Universal rule predicts, transcending geography, ethnicity and race.

The Universal rule requires that Herd immunity begin when just 25% of the population is vaccinated. With the anticipated 100 million vaccinations in the first 100 days of the Biden administration, Herd immunity may be imminent in the US.

The Universal rule promotes a temporary stasis with continuing infections and hospitalizations and becomes a barrier to runaway infections, making it practically impossible to reach Herd immunity, as Sweden discovered. Reduced infected population seems to be a third option to stifle the epidemic - a little known accomplishment, first by North Dakota and subsequently by twelve other U.S. states, including South Dakota.

## Introduction: Unusual challenges of the Covid 19 pandemic

The flood of global Covid 19 data, distributed daily by Johns Hopkins University (1), WHO (2) and other organizations around the world, can be both informative and intimidating. While the patient zero is not known, the biology of the Corona virus suggests that this is a single disease that has spread globally. But at a population level, the picture is baffling and less unified, exhibiting diverse spread and recovery rates of the disease. Studying patterns in global Covid 19 data seemed to be a good way to bring some clarity to this picture.

In particular, are Covid 19 data subject to hidden rules? Empirical rules in data, such as scaling or power and exponential laws, are hallmarks of “complex systems” (3 - 6). And should such rules be found, the lessons learned from critical phenomena could be adapted to the dynamics of epidemics. It is the time evolution of the pandemic that we seek to understand.

Then there is the Herd immunity puzzle. Because, infected people also become immune, some politicians and governments promoted mass infections as an alternate route to build immunity, so as to stop the spread without stalling the economy. But the disastrous experience in Sweden, where the pandemic spread, without Government restraint, was disconcerting. Thousands more died than in neighboring Nordic countries, but infections remained stubbornly at 7% in Sweden, far below the level needed for herd immunity. But the experience of others, like the passengers in the closed environment of the cruise ship Diamond Princess and amongst devotees during the mass religious gatherings in Iran have been different. What prevents runaway infections in this pandemic?

With mass vaccinations beginning around the world, there is an expectation for an alternate and more legitimate path to real Herd immunity soon. But the estimates of when that will happen are wildly different, ranging from 60% to over 80% of total population being vaccinated. Worse, the estimates are provided in the spirit of the Delphic oracle, without any explanation and attribution.

Since vast amounts of data are added daily, the search for rules by trial and error was not practical. Models are powerful tools to explore the dynamics of pandemics. True, the reputations of models have taken a hit during the Covid 19 pandemic but, as discussed later in this paper, modelers and the communities that use them must share blame, not the models.

The epidemic model chosen was the SIR model, published in the Proceedings of the Royal Society almost 94 years ago by Kermack and McKendrick (7). The Institute for Advanced Study at Princeton has a fascinating seal that memorializes “Truth and Beauty” in science, and the quote is from John Keats’ famous poem *Ode on a Grecian Urn*. But for the SIR model, the paraphrase of a popular saying attributed to ancient Indian sages, “When there is Satyam and Sundaram, Shivam is sure to follow” is more apt.

A Google search reveals that over 1.5 million papers have used the model, but, as we will find in this paper, there is much left to be discovered. Almost all of the papers use numerical and approximate solutions because the equations are difficult to solve exactly. In the process, the beauty and elegance of the model proves elusive. Our prior success with a modified SIR model, in a forensic problem involving Medicaid programs, exposed a technical quirk in the model and showed that slight modifications could yield unexpected and new results that are easy to analyze and interpret. It is ironic that we have come full circle, first, modifying the epidemic model to solve a forensic problem, and now, adapting the forensic model to understand a global pandemic.

## Methods

This section includes three mathematical results that are crucial to the paper. The first is the derivation of an exact solution to the 94-year-old SIR model. Next, the exact solution is used to reveal a hidden Universal rule, applicable to global Covid 19 data. The third is to derive the mathematical restrictions on the development of the epidemic placed by the Universal rule. While the derivations of these results are complete and mathematically rigorous, they are quite unnecessary to understand the validation and implication of the results. Once the results are accepted, they shed light on the failed experience in Sweden, the little-known success in controlling the epidemic in North and South Dakota and several other states, and how the 100 million vaccinations in the first hundred days is an implicit guarantee of Herd immunity for the entire nation.

### a. Criteria to select the ideal model

Physical systems, like the atmosphere and living organisms are far too complex to describe exactly using mathematics. The goal is to create a simple solvable model that captures the essence of the complex system, so as to obtain a deeper understanding of the whole. Often, they are only partially correct in replicating a process. But they can still yield important insight.

When it comes to epidemics, the SIR model (8 - 10) belongs to a class known as “compartmental models,” and is most popular. But there is a hitch. The model is simple to describe, but the equations of the model are too difficult to solve exactly. The central issue is a technical one. The equations of the model are nonlinear, and these are difficult to handle mathematically, even when they appear to be deceptively simple. Some of these challenges are described in Hirsch et al. (11) and Strogatz (12).

Simple equations leading to complex science is not unique to epidemiology. For example, the entire subject of “chaos” owes its origin to a set of nonlinear equations in an absurdly simple one-dimensional model (13) of the atmosphere. Similarly, Robert May, R.M. Anderson and others (14-16), analyzed the simple nonlinear predator prey equations to discover chaos lurking in biological systems.

### b. Epidemics to Medicaid and back to epidemics

Earlier, in dealing with chronic Medicaid populations, a modification to the SIR model made the equations solvable, making it possible to forecast prevalence, incidence and costs of treatment of chronic populations with great accuracy. Simply put, the exact nonlinear equations that cannot be solved were replaced with approximate linear equations that can be solved exactly.

This linearization yielded a disease and cost forecasting model for chronic and cancer patients. The cost forecasts showed a national pattern of Medicaid overpayments, estimated to be in excess of $100 billion. Validation came in the form of recovery of funds. Specifically, the forecasts detected $250 million in overpayments to health insurers in 2014, in one state. The state initially disputed this finding because its annual audits missed the overpayments. But in a report (17) to the legislature in 2019, the state affirms that the overpayment pattern discovered through the SIR analyses continued until 2017, and the state had recovered $660 million in total.

Since it was the modification of the epidemic model that helped address the “endemic” Medicaid cost issue in one state, it made sense, in a perverse way, to try and modify the Medicaid solution to solve the Covid 19 puzzle. Pursuing a similar analysis, it soon became obvious that linearization for Medicaid worked only because physical contact expressed through the nonlinear terms is unimportant in the spread of chronic diseases and cancer. For Covid 19, physical contact is critical, and nonlinear terms cannot be ignored. But there was hope. Hirsch et al., on page 295 of their book on dynamical systems (11) suggest “rescaling” as an alternate mathematical technique that in some cases convert unsolvable nonlinear equations to solvable linear ones. Surprisingly, this worked, as described below.

### c. The exact solution of the SIR model

For those readers not mathematically inclined, we first provide a verbal description of the model, its technical challenges and the different approach we adopted to reach a solution. We cannot ignore the mathematics, because it is the rules of mathematics that sunk Sweden’s “FrankenHerd” experiment and the same rules that promise Herd immunity for the nation in the first hundred days of the Biden administration.

In the SIR model, population is segmented into three compartments – those who are Susceptible (*S*), those Infected *(I)* and the third called Removed *(R)*, consisting of those who recovered or died. As time passes, people in one compartment transition to another. There are two parameters in the model, *β* to describe the “infection” rate and *γ* for the “removal” (a group that is essentially immunized) rate. One of the equations deals with the balance between infection and removals and, in effect, describes the progress of the epidemic. Mathematically speaking, it is the “sign” of this equation — positive for increase and negative for dwindling of the infected population — that determines whether the epidemic is growing or waning.

There are three distinct ways in which this can happen. First, reducing the infected population; second, increasing the removed population and thus making more people immune to the disease, as Sweden tried; and the third, vaccination to reduce the number of susceptible people by immunizing many. The second and third options are called Herd immunity but not the first option, even though it also results in damping the growth of the epidemic.

There are product terms (*I S)* in the equations and those are the nonlinear terms that make it difficult to explicitly solve the equations of the SIR model. We used a mathematical technique called “rescaling” to get rid of the nonlinear terms so as to obtain an exact solution. Those uninterested in the mathematical derivation of the exact solution, the derivation of the Universal rule and why the Herd immunity conditions are mathematically dependent on the Universal rule, can skip to the “Results” section.

The three equations describing the time evolution of the three population segments are:

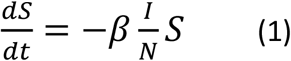

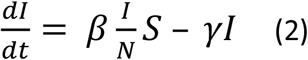

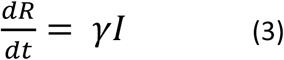

Both *β* and *γ* are parameters of the model. As defined above, *β I* is the rate of infection while *γ* is the rate of removal. *N* is the total population, defined as

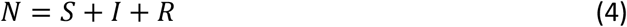

Since the infection and removal periods are short, the total number of people in the population is assumed to be a constant. This constancy is built into the equations by ensuring that the sum of eqs. (1), (2) and (3) adds to zero.

We also assume that at time zero, almost the entire population is Susceptible.

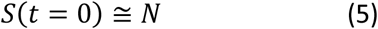

While the model is very simple to describe, the nonlinear product term (I S) in eqs. (1) and (2) make them difficult to solve. Almost always the only solutions available are numerical, and they do not lend themselves to analysis, or to the discovery of the new results derived in the paper.

“Rescaling” refers to defining a new independent variable to rescale time ‘t’ to a pseudo time variable defined as

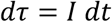

And then the modified equations become linear.

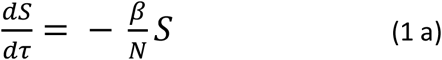

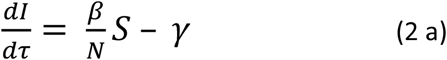

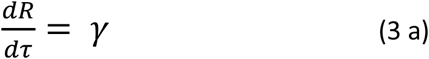

The new pseudo-time variable *τ* is defined as a definite integral

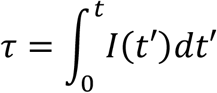

Since I(t) is included in the Johns Hopkins data on a daily basis, the integral can be calculated easily using a program like MATLAB or even using the trapezoidal rule.

Because of the constraint equation (4), only two out of the three equations are independent. Most importantly, because the rescaled equations are linear, these equations can be integrated immediately, to obtain exact analytic solutions that are easy to differentiate and interpret.

From (1 a) we get a simple exponential solution:

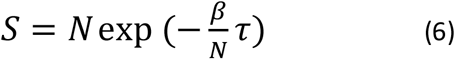

where we have used the boundary condition (5) to calculate the constant of integration.

Similarly, from eq (3a) we get

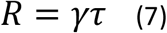

And using (4) we have an expression for *I*.

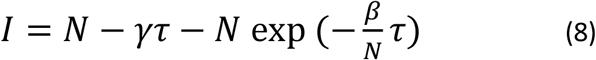

Equations (6), (7) and (8) provide a complete, exact analytical solution for the SIR model. In particular, we can use the data to calculate the values of the parameters, *β* and *γ*.

From eq. (6) we get

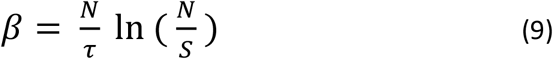

And from eq. (7)

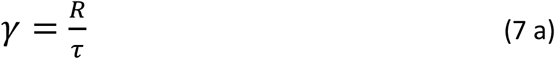

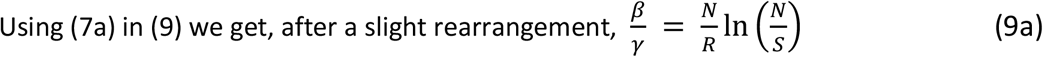

While the derivations are important, it is the exact and analytical results in the form of eqs. (6,7, 8, 9, 7a and 9a) that is crucial to appreciate the advances described in this paper.

A side observation is that eq. (9a) can also be derived from the original equations, without rescaling. Page 235 of Hirsch et al. (11), contains an expression for I(S) that, when integrated with the appropriate boundary condition leads to eq. (9a) without using the exact solution. But, the analytic solution is required to derive eq (9).

### d. *Universal* rule revealed

In physics, Emmy Noether’s elegant theorem (18) connects symmetries in data to constraint equations. That prompted the analysis of our constraint equation, using our analytic solution.

The constraint eq. (4) can be rewritten as

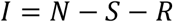

Substituting for *S*, using eq. (6), and expanding the exponential in a power series and retaining just the term linear in *τ*, leads to a version of the universal rule

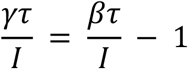

and it describes a line, relating the quantities 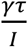 and 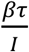 with slope equal to 1 and Y-intercept equal to (−1). It is very important to remember that this is an approximate result obtained by retaining only the first order terms in *τ*.

And we are using the analytic solution and the Universal rule in a crucial way to derive this expression. In other words, numerical approximations will not reveal the Universal rule. It needs the exact analytic solution.

It is possible to eliminate *τ* by using (7a), and rewriting the equation as

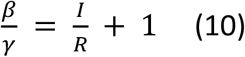

eq. (10) is the version of the Universal rule we will use. It is a rule that is startling in its simplicity: 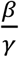 that measures ratio of infection and removal rates is linearly related to ratios of observables (or measurable quantities) I and R. An alternate way of describing this equation is that 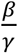 and 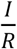, when graphed will fall on a line, and they move in lock step, perfectly correlated.

The conclusion is that, *if the SIR model is a valid description* of the Covid 19 pandemic, then the Universal rule must apply for the entire set of global Covid 19 data. It is a very restrictive rule. The datapoints 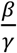, I and R for any country or state, and for any date, *must all fall on the same line* with slope and Y intercept, both equal to 1. If the data fail to match the rule, and bereft of an adequate explanation, it is time to consider a different model.

### e. Herd Immunity

From a mathematical standpoint, Herd immunity begins when the epidemic begins to decay, when rate of change of infected population decreases with time. In effect, the number removed (those recovered and dead) is greater than the number infected, and so, the size of the infected population shrinks as time passes. The relevant equation is (2a) where the change in the infected (I), 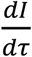 becomes negative or

when, 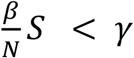 in eq. (2a) or equivalently

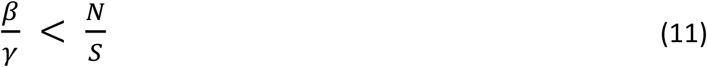

But the Universal rule eq (10), which also involves 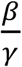 imposes restrictions on S in eq. (11).

Rewriting eq. (10) and using constraint equation (4),

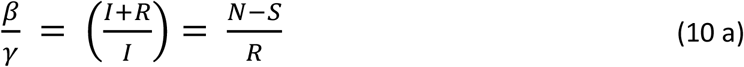

From the RHS of (10 a) and (11) we get the inequality as a quadratic inequality 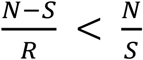 and this can be expressed as a quadratic inequality

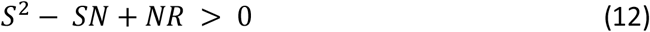

In other words, for the epidemic to dissipate, S must satisfy this quadratic inequality. The following analysis will yield three distinct paths to pause the growth of the epidemic, two of them are called Herd immunity.

Elementary algebra texts describe the derivation of solutions to a quadratic inequality. Google search yields many options. Finding the solution starts with solving a related equation where the RHS of eq (12) is set to zero.

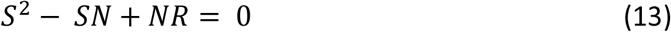

The first step is to find the roots of this modified quadratic “equation.” The roots can be real or complex, and they have to be handled in slightly different ways. The idea is to use the roots of the equation (13) to find the range of real values of S that satisfy the inequality eq. (12)

The two roots *S*_1_ and *S*_2_ that satisfy eq. (13) are:

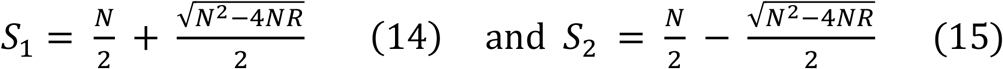

eq. (14) and eq. (15) will help find those values of S that satisfy the inequality eq. (12)

There are two cases to consider and they will lead to three paths to damp the growth of the epidemic.

Case (a): Real roots when *N*^2^ *−* 4*NR* is not negative, or equivalently, 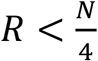

The roots *S*_1_ and *S*_2_, are located where the curve defined by eq. (12) intersects the X-axis (S is along X axis). Now it is possible to identify the regions where the S values satisfy the inequality and thus offer two paths to stifle the growth of the epidemic:

*S* > *S*_1_ and *S* < *S*_2_, both while 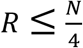.

Case (b): Complex roots, when *N*^2^ *−* 4*NR* is negative, or when, 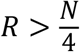

It is tempting to ignore complex roots since we are dealing with a problem with real numbers. But that would be a serious omission, as we discovered.

It is important to remember that our interest is in finding the range of values of S that will satisfy the inequality eq. (12) and NOT the solution for eq. (13). So, the lack of a valid real solution for eq. (13) is immaterial.

In the case of complex roots, the S curve defined in eq. (12) does not intersect the X axis. Now, as before, we have to find the values of S that can still satisfy eq. (12). It turns out that every *allowed value of S* (positive and real numbers) satisfies the inequality in eq (12). Thus, the third path to stifling the epidemic is to increase R, at will, through vaccination.

In summary, *S* > *S*_1_ while 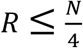 stops the growth of the epidemic by lowering infections but is not classified as Herd immunity. *S* < *S*_2_ while 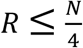 is the Herd immunity that Sweden failed to achieve. And 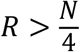 is the Herd immunity reached through vaccination.

From (9a) and (11) we can derive

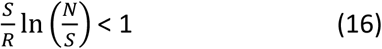

and it defines the Herd immunity condition for any population.

### f. Source of data

All the data used in this paper came from “COVID-19 Data Repository by the Center for Systems Science and Engineering (CSSE) at Johns Hopkins University.” The GitHub repository (19) permits the download of the data in a CVS format.

Since we are plotting 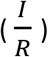 we cannot include data points where R in the denominator is zero. All states, except for Wyoming, have R greater than zero, starting from April 1. Wyoming has R greater than 0 after April 15 2020. The period we chose ends on December 12, 2020 just before the vaccines were introduced to the US population. For the international countries also we adopted April 1 as a starting date. About 10 countries (mostly small islands) had R equal to zero and very low infection rates.

### g. Calculation of best linear fits to data

Excel and Tableau were used to plot the various graphs and also to calculate the relevant trendlines. Tableau offers an unparalleled dynamic visual rendering of the Universal rule as a time series, and can be used to compare the data of states and countries easily. Since R =0 at the start, 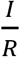 is usually large in the beginning and then moving to the origin as time progresses. Tableau captures this time series vividly.

For a quantitative but numerical view of the data, we used Excel and its trendlines. The first trendline is just the least squares fit using multiple linear regression. Since the Universal rule as a constant Y-intercept of unity, it also made sense to have Excel generate a second trendline that has a Y-intercept of 1.

All the states of the US and 50 of the countries with the most confirmed cases are listed in the Tables 1 and 2.

**Table 1.**
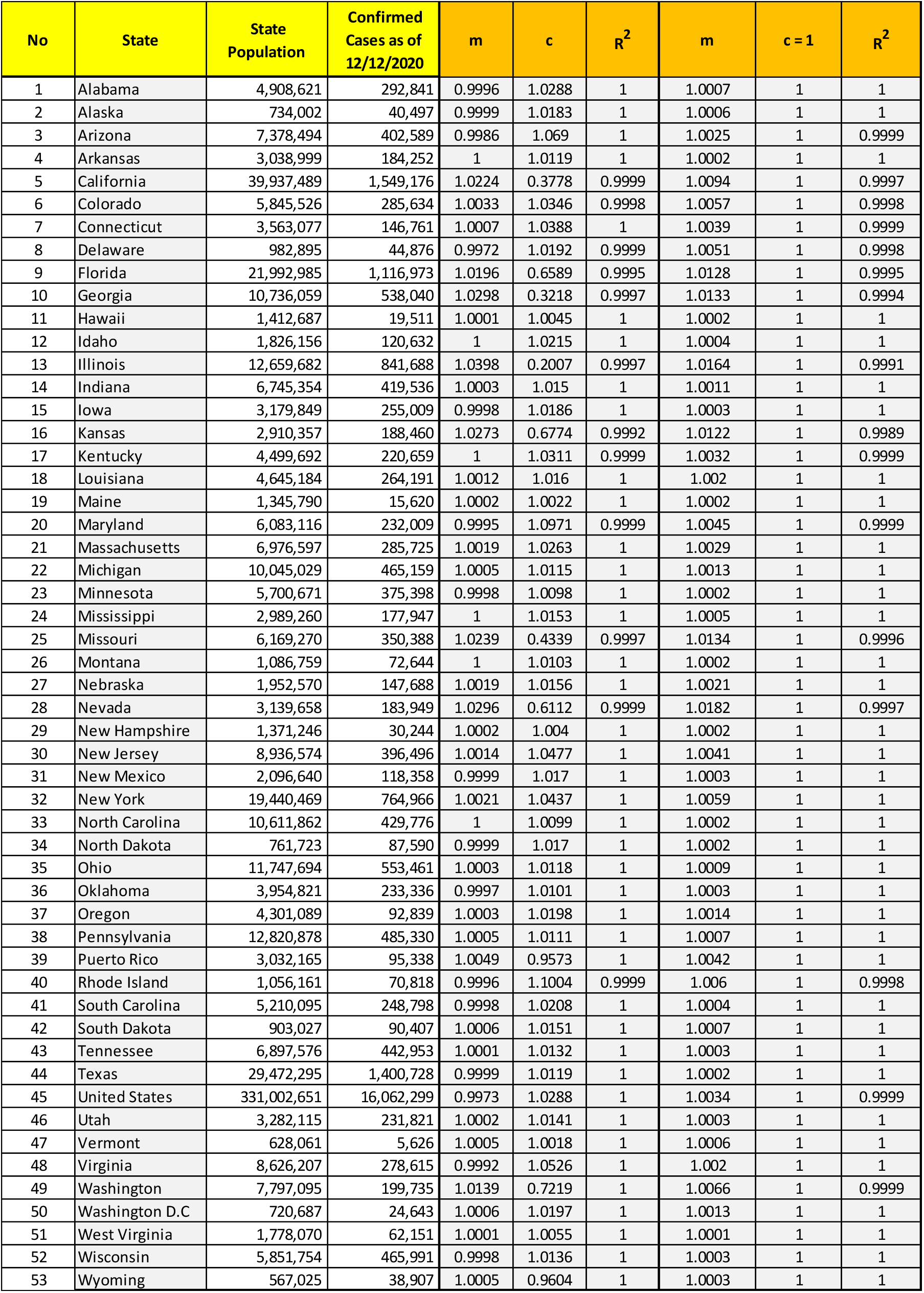
US states and the Universal rule. Line parameters Slope (m), intercept (c) and **R**^**2**^ (correlation coefficient) for least squares fit and for a set where Y-intercept is fixed as 1.

| No | State | State Population | Confirmed Cases as of 12/12/2020 | m | c | R<sup>2</sup> | m | c = 1 | R<sup>2</sup> |
| --- | --- | --- | --- | --- | --- | --- | --- | --- | --- |
| 1 | Alabama | 4,908,621 | 292,841 | 0.9996 | 1.0288 | 1 | 1.0007 | 1 | 1 |
| 2 | Alaska | 734,002 | 40,497 | 0.9999 | 1.0183 | 1 | 1.0006 | 1 | 1 |
| 3 | Arizona | 7,378,494 | 402,589 | 0.9986 | 1.069 | 1 | 1.0025 | 1 | 0.9999 |
| 4 | Arkansas | 3,038,999 | 184,252 | 1 | 1.0119 | 1 | 1.0002 | 1 | 1 |
| 5 | California | 39,937,489 | 1,549,176 | 1.0224 | 0.3778 | 0.9999 | 1.0094 | 1 | 0.9997 |
| 6 | Colorado | 5,845,526 | 285,634 | 1.0033 | 1.0346 | 0.9998 | 1.0057 | 1 | 0.9998 |
| 7 | Connecticut | 3,563,077 | 146,761 | 1.0007 | 1.0388 | 1 | 1.0039 | 1 | 0.9999 |
| 8 | Delaware | 982,895 | 44,876 | 0.9972 | 1.0192 | 0.9999 | 1.0051 | 1 | 0.9998 |
| 9 | Florida | 21,992,985 | 1,116,973 | 1.0196 | 0.6589 | 0.9995 | 1.0128 | 1 | 0.9995 |
| 10 | Georgia | 10,736,059 | 538,040 | 1.0298 | 0.3218 | 0.9997 | 1.0133 | 1 | 0.9994 |
| 11 | Hawaii | 1,412,687 | 19,511 | 1.0001 | 1.0045 | 1 | 1.0002 | 1 | 1 |
| 12 | Idaho | 1,826,156 | 120,632 | 1 | 1.0215 | 1 | 1.0004 | 1 | 1 |
| 13 | Illinois | 12,659,682 | 841,688 | 1.0398 | 0.2007 | 0.9997 | 1.0164 | 1 | 0.9991 |
| 14 | Indiana | 6,745,354 | 419,536 | 1.0003 | 1.015 | 1 | 1.0011 | 1 | 1 |
| 15 | Iowa | 3,179,849 | 255,009 | 0.9998 | 1.0186 | 1 | 1.0003 | 1 | 1 |
| 16 | Kansas | 2,910,357 | 188,460 | 1.0273 | 0.6774 | 0.9992 | 1.0122 | 1 | 0.9989 |
| 17 | Kentucky | 4,499,692 | 220,659 | 1 | 1.0311 | 0.9999 | 1.0032 | 1 | 0.9999 |
| 18 | Louisiana | 4,645,184 | 264,191 | 1.0012 | 1.016 | 1 | 1.002 | 1 | 1 |
| 19 | Maine | 1,345,790 | 15,620 | 1.0002 | 1.0022 | 1 | 1.0002 | 1 | 1 |
| 20 | Maryland | 6,083,116 | 232,009 | 0.9995 | 1.0971 | 0.9999 | 1.0045 | 1 | 0.9999 |
| 21 | Massachusetts | 6,976,597 | 285,725 | 1.0019 | 1.0263 | 1 | 1.0029 | 1 | 1 |
| 22 | Michigan | 10,045,029 | 465,159 | 1.0005 | 1.0115 | 1 | 1.0013 | 1 | 1 |
| 23 | Minnesota | 5,700,671 | 375,398 | 0.9998 | 1.0098 | 1 | 1.0002 | 1 | 1 |
| 24 | Mississippi | 2,989,260 | 177,947 | 1 | 1.0153 | 1 | 1.0005 | 1 | 1 |
| 25 | Missouri | 6,169,270 | 350,388 | 1.0239 | 0.4339 | 0.9997 | 1.0134 | 1 | 0.9996 |
| 26 | Montana | 1,086,759 | 72,644 | 1 | 1.0103 | 1 | 1.0002 | 1 | 1 |
| 27 | Nebraska | 1,952,570 | 147,688 | 1.0019 | 1.0156 | 1 | 1.0021 | 1 | 1 |
| 28 | Nevada | 3,139,658 | 183,949 | 1.0296 | 0.6112 | 0.9999 | 1.0182 | 1 | 0.9997 |
| 29 | New Hampshire | 1,371,246 | 30,244 | 1.0002 | 1.004 | 1 | 1.0002 | 1 | 1 |
| 30 | New Jersey | 8,936,574 | 396,496 | 1.0014 | 1.0477 | 1 | 1.0041 | 1 | 1 |
| 31 | New Mexico | 2,096,640 | 118,358 | 0.9999 | 1.017 | 1 | 1.0003 | 1 | 1 |
| 32 | New York | 19,440,469 | 764,966 | 1.0021 | 1.0437 | 1 | 1.0059 | 1 | 1 |
| 33 | North Carolina | 10,611,862 | 429,776 | 1 | 1.0099 | 1 | 1.0002 | 1 | 1 |
| 34 | North Dakota | 761,723 | 87,590 | 0.9999 | 1.017 | 1 | 1.0002 | 1 | 1 |
| 35 | Ohio | 11,747,694 | 553,461 | 1.0003 | 1.0118 | 1 | 1.0009 | 1 | 1 |
| 36 | Oklahoma | 3,954,821 | 233,336 | 0.9997 | 1.0101 | 1 | 1.0003 | 1 | 1 |
| 37 | Oregon | 4,301,089 | 92,839 | 1.0003 | 1.0198 | 1 | 1.0014 | 1 | 1 |
| 38 | Pennsylvania | 12,820,878 | 485,330 | 1.0005 | 1.0111 | 1 | 1.0007 | 1 | 1 |
| 39 | Puerto Rico | 3,032,165 | 95,338 | 1.0049 | 0.9573 | 1 | 1.0042 | 1 | 1 |
| 40 | Rhode Island | 1,056,161 | 70,818 | 0.9996 | 1.1004 | 0.9999 | 1.006 | 1 | 0.9998 |
| 41 | South Carolina | 5,210,095 | 248,798 | 0.9998 | 1.0208 | 1 | 1.0004 | 1 | 1 |
| 42 | South Dakota | 903,027 | 90,407 | 1.0006 | 1.0151 | 1 | 1.0007 | 1 | 1 |
| 43 | Tennessee | 6,897,576 | 442,953 | 1.0001 | 1.0132 | 1 | 1.0003 | 1 | 1 |
| 44 | Texas | 29,472,295 | 1,400,728 | 0.9999 | 1.0119 | 1 | 1.0002 | 1 | 1 |
| 45 | United States | 331,002,651 | 16,062,299 | 0.9973 | 1.0288 | 1 | 1.0034 | 1 | 0.9999 |
| 46 | Utah | 3,282,115 | 231,821 | 1.0002 | 1.0141 | 1 | 1.0003 | 1 | 1 |
| 47 | Vermont | 628,061 | 5,626 | 1.0005 | 1.0018 | 1 | 1.0006 | 1 | 1 |
| 48 | Virginia | 8,626,207 | 278,615 | 0.9992 | 1.0526 | 1 | 1.002 | 1 | 1 |
| 49 | Washington | 7,797,095 | 199,735 | 1.0139 | 0.7219 | 1 | 1.0066 | 1 | 0.9999 |
| 50 | Washington D.C | 720,687 | 24,643 | 1.0006 | 1.0197 | 1 | 1.0013 | 1 | 1 |
| 51 | West Virginia | 1,778,070 | 62,151 | 1.0001 | 1.0055 | 1 | 1.0001 | 1 | 1 |
| 52 | Wisconsin | 5,851,754 | 465,991 | 0.9998 | 1.0136 | 1 | 1.0003 | 1 | 1 |
| 53 | Wyoming | 567,025 | 38,907 | 1.0005 | 0.9604 | 1 | 1.0003 | 1 | 1 |

**Table 2:** Top 50 Countries with most confirmed cases and the Universal rule: Line parameters Slope (m), intercept (c) and **R**^**2**^ (correlation coefficient) for least squares fit and for a set where Y-intercept is fixed as 1.

| Country | Country Population | Confirmed Cases as of 12/12/2020 | m | c | R<sup>2</sup> | m | c=1 | R<sup>2</sup> |
| --- | --- | --- | --- | --- | --- | --- | --- | --- |
| US | 331,002,651 | 16,062,299 | 0.9974 | 1.0285 | 1 | 1.0035 | 1 | 0.9999 |
| India | 1,380,004,385 | 9,857,029 | 0.9996 | 1.0019 | 1 | 1.0001 | 1 | 1 |
| Brazil | 211,049,527 | 6,880,127 | 0.9993 | 1.0091 | 1 | 1.0002 | 1 | 1 |
| Russia | 145,872,256 | 2,602,048 | 0.9996 | 1.0054 | 1 | 1.0003 | 1 | 1 |
| France | 65,273,511 | 2,405,255 | 1.0191 | 0.9755 | 1 | 1.0151 | 1 | 0.9999 |
| United Kingdom | 67,886,011 | 1,835,949 | 1.0143 | 0.9205 | 1 | 1.0089 | 1 | 0.9999 |
| Italy | 60,461,826 | 1,825,775 | 1.0058 | 1.0032 | 0.9999 | 1.0083 | 1 | 0.9999 |
| Turkey | 84,339,067 | 1,809,809 | 1.0001 | 1.0023 | 1 | 1.0003 | 1 | 1 |
| Spain | 46,754,778 | 1,730,575 | 1.0196 | 0.9884 | 1 | 1.0172 | 1 | 1 |
| Argentina | 44,780,677 | 1,494,602 | 0.9942 | 1.0128 | 1 | 1.0011 | 1 | 0.9999 |
| Colombia | 50,339,443 | 1,417,072 | 1.0049 | 1 | 0.9999 | 1.0049 | 1 | 0.9999 |
| Germany | 83,783,942 | 1,336,101 | 1.0013 | 1.0023 | 1 | 1.0037 | 1 | 0.9999 |
| Mexico | 127,575,529 | 1,241,436 | 0.9998 | 1.0022 | 1 | 1.0001 | 1 | 1 |
| Poland | 37,846,611 | 1,126,700 | 0.9998 | 1.00E+00 | 1 | 1.0003 | 1 | 1 |
| Iran | 83,992,949 | 1,100,818 | 0.9999 | 1.0027 | 0.9999 | 1.0058 | 1 | 0.9998 |
| Peru | 32,510,453 | 980,943 | 0.9934 | 1.0138 | 1 | 1.0033 | 1 | 0.9998 |
| Ukraine | 43,993,638 | 908,839 | 0.9997 | 1.0054 | 1 | 1.0002 | 1 | 1 |
| South Africa | 58,558,270 | 852,965 | 0.9996 | 1.0047 | 1 | 1.0001 | 1 | 1 |
| Netherlands | 17,134,872 | 613,630 | 1.019 | 0.8627 | 1 | 1.0116 | 1 | 0.9999 |
| Indonesia | 270,625,568 | 611,631 | 0.9998 | 1.0007 | 1 | 1.0001 | 1 | 1 |
| Belgium | 11,589,623 | 603,159 | 1.0271 | 0.9562 | 1 | 1.0252 | 1 | 1 |
| Czechia | 10,708,981 | 575,422 | 0.9999 | 1.0088 | 1 | 1.0004 | 1 | 1 |
| Iraq | 39,309,783 | 573,622 | 0.9947 | 1.0057 | 1 | 1.002 | 1 | 0.9999 |
| Chile | 18,952,038 | 569,781 | 0.9981 | 1.0103 | 1 | 1.0009 | 1 | 0.9999 |
| Romania | 19,237,691 | 551,900 | 0.9989 | 1.0055 | 1 | 1.001 | 1 | 1 |
| Bangladesh | 163,046,161 | 489,178 | 0.9999 | 1.0013 | 1 | 1 | 1 | 1 |
| Canada | 37,971,020 | 458,527 | 0.9994 | 1.0028 | 1 | 1.0009 | 1 | 1 |
| Philippines | 108,116,615 | 448,331 | 0.9998 | 1.0014 | 1 | 1.0001 | 1 | 1 |
| Pakistan | 216,565,318 | 438,425 | 0.9999 | 1.0008 | 1 | 1.0001 | 1 | 1 |
| Morocco | 36,471,769 | 397,597 | 0.9996 | 1.0019 | 1 | 1.0001 | 1 | 1 |
| Switzerland | 8,591,365 | 373,831 | 1.0073 | 1.0047 | 0.9997 | 1.0115 | 1 | 0.9997 |
| Saudi Arabia | 34,268,528 | 359,749 | 0.9993 | 1.0046 | 1 | 1.0004 | 1 | 1 |
| Israel | 8,519,377 | 355,786 | 0.9995 | 1.0104 | 1 | 1.0008 | 1 | 1 |
| Portugal | 10,196,709 | 344,700 | 1.0004 | 1.0071 | 1 | 1.0008 | 1 | 1 |
| Sweden | 10,099,265 | 320,098 | 1.0146 | 0.8893 | 1 | 1.0091 | 1 | 0.9999 |
| Austria | 8,955,102 | 319,822 | 1.0011 | 1.0042 | 0.9999 | 1.0037 | 1 | 0.9999 |
| Hungary | 9,660,351 | 276,247 | 1.0023 | 1.0029 | 0.9999 | 1.0032 | 1 | 0.9999 |
| Serbia | 8,772,235 | 261,437 | 1.0085 | 0.8533 | 1 | 1.0063 | 1 | 1 |
| Jordan | 10,101,694 | 257,275 | 1.0055 | 0.9987 | 1 | 1.0053 | 1 | 1 |
| Nepal | 28,608,710 | 247,593 | 0.9997 | 1.0022 | 1 | 1.0001 | 1 | 1 |
| Ecuador | 17,373,662 | 201,524 | 0.9999 | 1.0041 | 1 | 1.0005 | 1 | 1 |
| Panama | 4,246,439 | 190,585 | 0.9998 | 1.0131 | 1 | 1.0004 | 1 | 1 |
| Georgia | 3,996,765 | 187,006 | 0.9986 | 1.0046 | 1 | 1.0011 | 1 | 0.9999 |
| Kazakhstan | 18,551,427 | 185,513 | 0.9997 | 1.0028 | 1 | 1.0001 | 1 | 1 |
| United Arab Emirates | 9,770,529 | 183,755 | 0.9995 | 1.0052 | 1 | 1.0004 | 1 | 1 |
| Bulgaria | 6,948,445 | 178,952 | 0.9999 | 1.0051 | 1 | 1.0013 | 1 | 1 |
| Japan | 126,860,301 | 178,272 | 1 | 1.0003 | 1 | 1.0001 | 1 | 1 |
| Croatia | 4,105,267 | 172,523 | 0.9996 | 1.004 | 1 | 1.0005 | 1 | 1 |

**Table 3.**
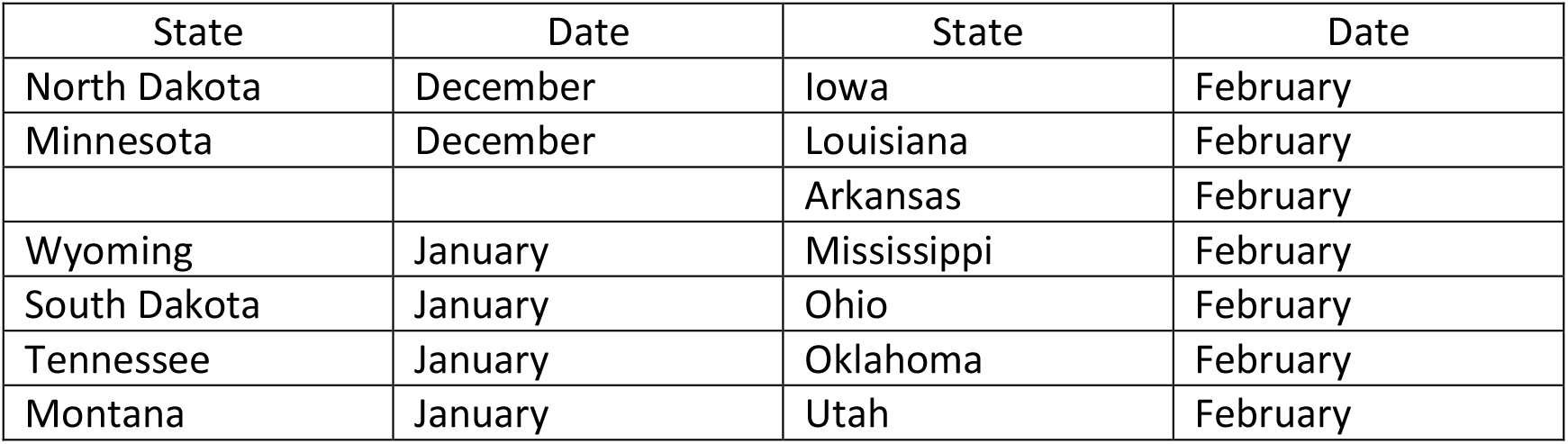
States and onset dates when the epidemic stopped growing

### h. Supplemental files

The Excel files with the entire set of global data used in the paper is included as supplemental files. This includes the entire set of 190 international countries as well as the states of the US, plus DC and Puerto Rico. The data are from Johns Hopkins.

The scatter graphs for every state and country are also included in the Excel data files.

## Results

In this section we will first check the validity of the Universal rule using global Covid 19 data. After confirmation of the Universal rule, we will examine its implications on the growth and dampening of the epidemic, as well as its impact on the two types of Herd immunity.

### (i) Validation of the Universal Rule using global Covid 19 data

Until this point, all of this is just theory – based on mathematics. In this section we will validate the Universal rule, using available data, to graphically prove the linear relationship between 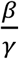 and 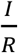 as in eq. (10)

This analysis includes the data published by the Johns Hopkins University (19), for the period April 1 to December 12, 2020, for all the states in the US (as well as data from Puerto Rico and DC) and 190 countries of the world. The plot is of 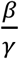 (on Y axis) and 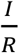 (on X axis). There is a total of over 186,000 data points. Remarkably, as the theory predicts, all the data fall on one line with slope 1 and Y intercept equal to 1. (These line graphs are of a time series obtained using Tableau.)

The graphs for other states and countries look exactly the same as Fig. 1, except that we have different ranges of values for 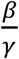 and 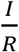. Fig. 2 and Fig. 3 are illustrative samples, one for the state of New Mexico and the other for Germany, and in both cases the data points fall on a line with slope 1 and Y-intercept 1, confirming the validity of the Universal rule.

**Fig 1.**
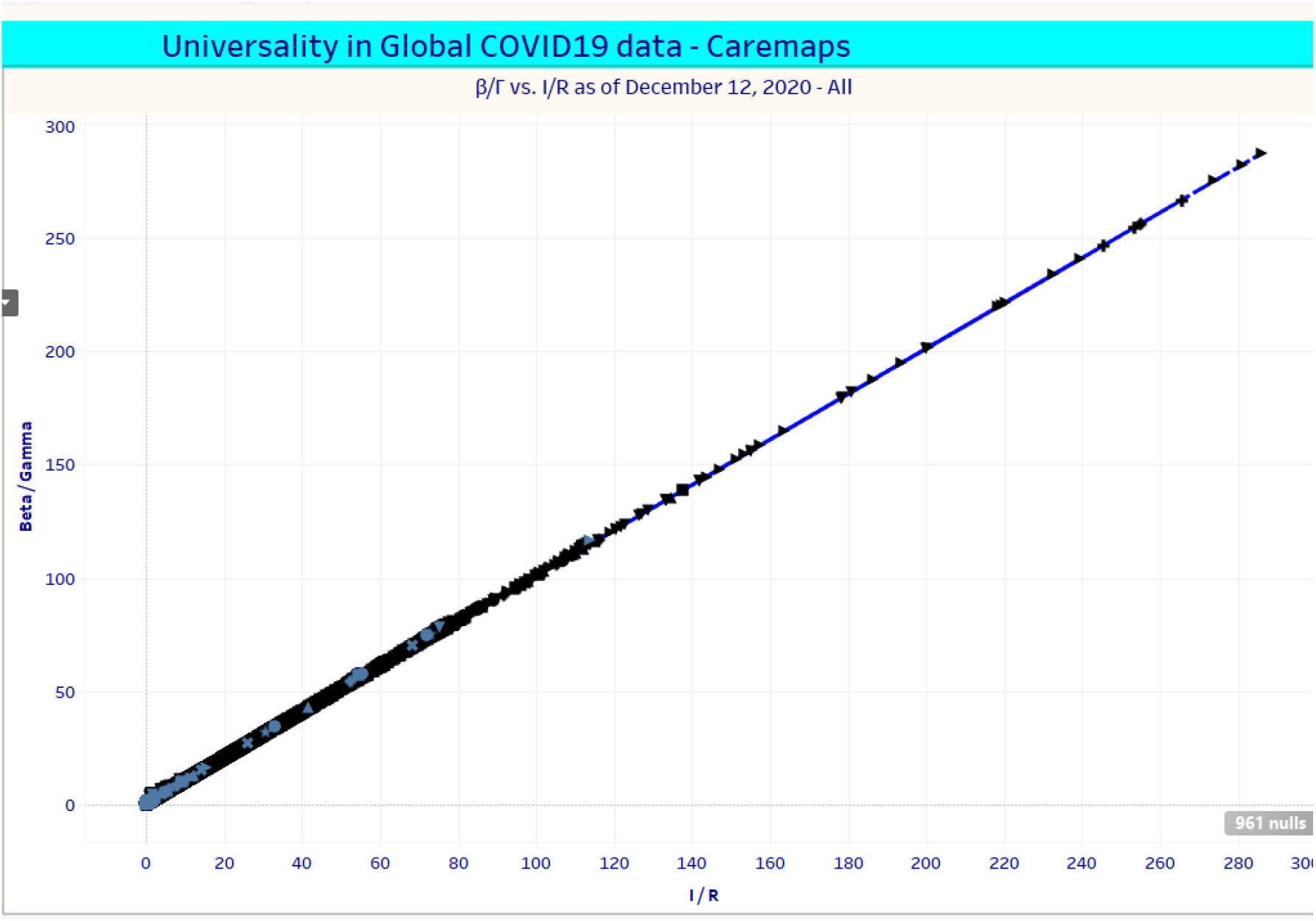
Global Covid 19 data conform to the universal rule. The points are from data for the period April 1 to December 12, 2020 from all the states in the US, and the 190 countries of the world. All data points fall on a single line, confirming the universal rule. (Screen shot from Tableau.)

**Fig 2.**
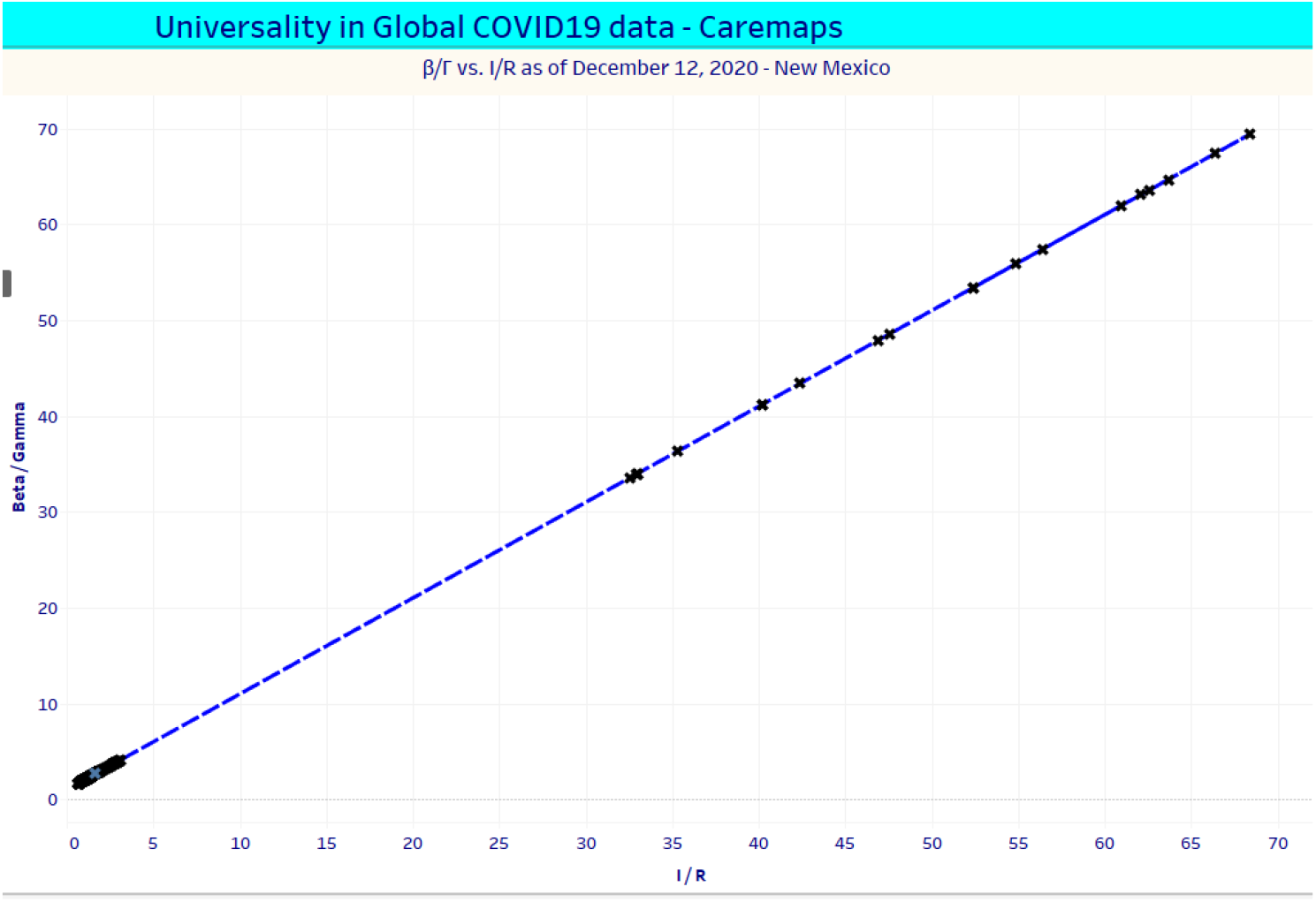
Universal rule for New Mexico. 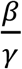 and 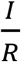 are for the period April 1 to December 12, 2020. Slope of the universal line is 1 and the Y-intercept 1. (Screenshot from Tableau.)

**Fig 3.**
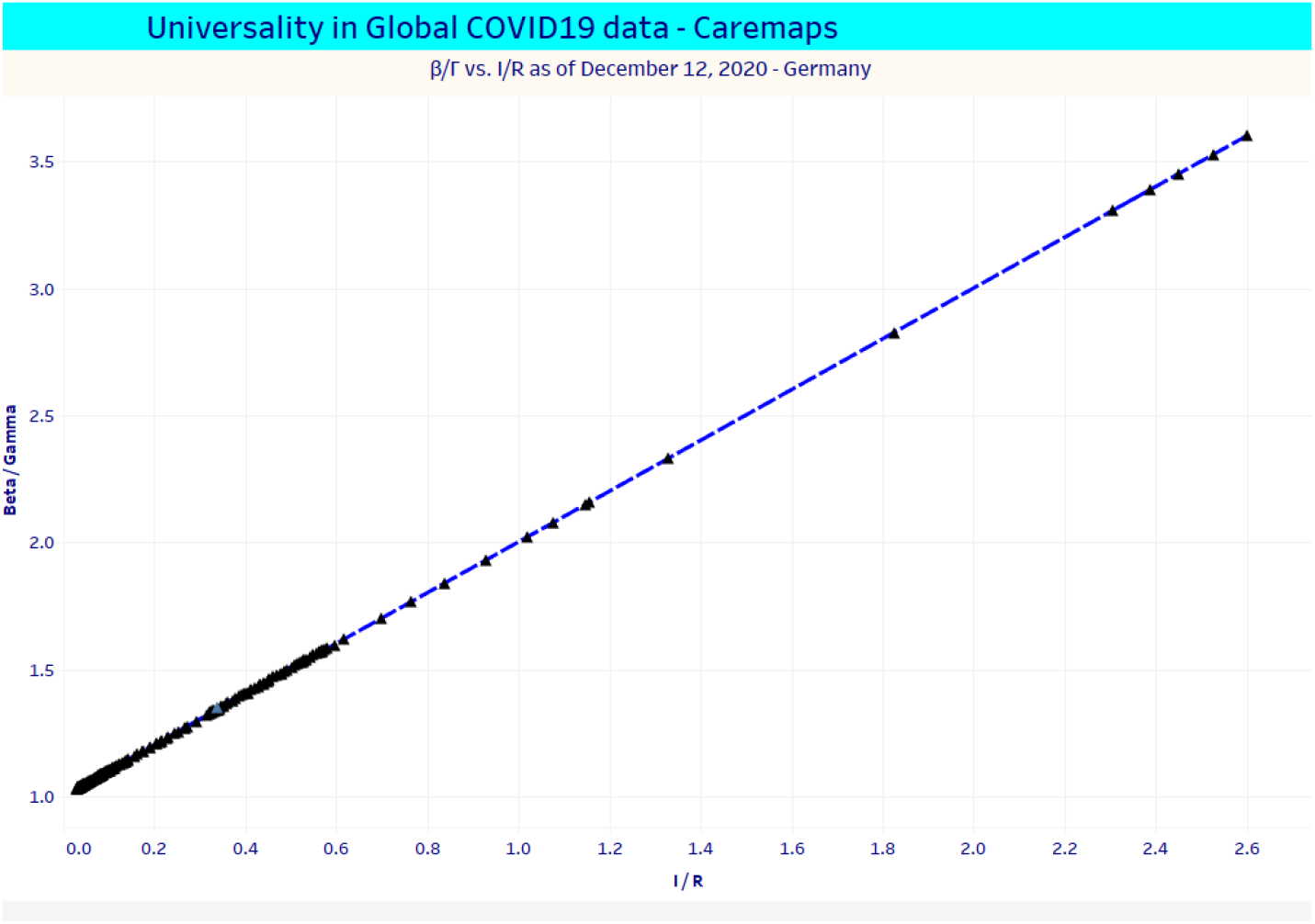
Universal rule for Germany. 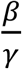 and 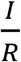 are for the period April 1 to December 12, 2020. The slope of the universal line is 1 and the Y-intercept 1. (Screenshot from Tableau.)

Even though the data points fall on a line, as prescribed by the universal rule, they are not ordered on the line sequentially in time. As time passes, the points can move up and down on the line irregularly, even while the values of 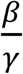 and 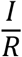 are perfectly correlated.

The graphs are all similar and it is very difficult to discern differences in them. Rather than provide over 200 similar looking graphs, we have created two tables, one for the nation’s states and the other for 50 international countries with the most confirmed cases.

First, we used Excel to plot the scatter graphs and create trendlines for each state or country, and obtained the equation for the trendline in terms of the slopes. Y-intercept values of the lines and their R^2^ values that measure correlations between 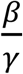 and 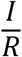. This is nothing more than the standard multiple linear regression calculations to obtain the best (least squares) fit. These are provided in Tables 1 and 2 below.

Excel also offers an option to fix the Y-intercept and then find the best fit of a trendline. Since Y-intercept has a special meaning in this analysis, as explained later, there is a second trendline calculated for each state and country and the slopes and the R^2^ values are displayed in the last three columns of Tables 1 and 2.

The Universal rule has been checked for 190 countries. For reasons of brevity, only 50 countries that have the most confirmed cases, as of Dec. 12, 2020, are listed in Table 2. The complete list and the supporting Excel files are in the attached supplement.

### (ii) Stopping the epidemic from growing

The term “herd immunity”, as used in public health is intended as a benign expression, to use vaccines to make a person immune to the disease while maintaining their health, and thus reduce its spread (20, 21). In many cases, people once infected, can develop immunity. In a macabre twist, politicians, governments and members of think tanks have promoted immunity through runaway infections of the virus and continue to call this inhumane program herd immunity. Of course, there is no way of knowing how long this immunity lasts nor if there are serious residual medical issues, such as coronary and pulmonary problems, for the recovered.

In the Methods section we showed that when the epidemic stops growing, S satisfies the inequality

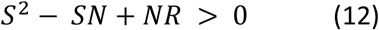

That is when 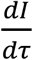 in eq. (2a) becomes negative, implying that removal rates are exceeding infection rates, and it is a signal for the start of the decay of the epidemic. There are three distinct ways the epidemic can stop growing.

#### (a) Controlling epidemic by lowering Infected population

When *S* > *S*_1_, and 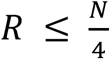, and if infections are low, then removals can exceed infections and the epidemic stops growing. This is not called Herd immunity but has the same end result.

Only one state, North Dakota, reached this stage as of December 12, 2020. More recent analysis identified a few other states where the epidemic stopped growing on later dates. Wyoming and Minnesota (in December); South Dakota, Tennessee and Montana (in January 2021); Iowa, Louisiana, Arkansas, Mississippi, Ohio, Oklahoma and Utah (in February, 2021).

#### (b) Controlling epidemic through runaway infections à la Sweden

The period where *S* < *S*_2_ is when the infections have grown to a substantial amount and there is reason to hope that Herd immunity can develop. (Again, there is an additional constraint 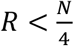 so as to ensure that *S*_2_ is real.) But the values of *S*_2_ in most cases is so small (maximum being less than 18% of total population), that the corresponding number of immunized becomes huge. S is even smaller, meaning over 82% of the total population must have been confirmed to be infected previously, in order to reach this limit.

The Swedish hope for Herd immunity belongs in this category. Unfortunately, this is highly improbable and practically speaking, impossible, because of the massive collateral damage inflicted from runaway infections. Sweden nor any other country ever reached these numbers of infections and Herd immunity remained beyond reach.

#### (d) Vaccinations and validations of Herd immunity

The third route to controlling the epidemic is the classic one, through vaccinations. Here, as deduced earlier, 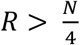, and all values of S satisfy the inequality and meet the threshold condition for Herd immunity.

In a typical compartmental model, it’s the infected (I) that moves to the removed (R) section, usually in about 30 days for Covid 19, and so one cannot increase without affecting the other. To permit increase in R without affecting the infected, we include transitions external to the model, in the form of vaccinations, and they allow susceptible populations S to transfer directly to the removed population R.

Eq. (16) defines the condition for reaching the stage when the epidemic stops growing. It is an inequality but all the quantities are observables, measured quantities supplied by Johns Hopkins, and so it is easy to verify if Herd immunity has been reached or if it is being sustained.

## Discussion

### Abuse of models in Covid 19 studies

Use of models has become a contentious issue during the Covid 19 pandemic. Part of the confusion is that the word model usually refers to a simplified representation of a natural process or a system, but the term has been hijacked, irresponsibly. A popular offender fits hospital utilization data to curves and calls it a model. That is just “curve fitting” which amounts to modeling a “result,” not a process. In guessing the shape of the curves describing hospital usage, for example, there is as much science as there is in picking tomorrow’s price of a volatile stock. Not stopping there, they extend the method to predict infection rates and mortality in the pandemic. Colorful phrases like “leveling the curve” emerge, but their predictions have been spectacularly wrong, sullying the reputation of real models.

For those who study or use models, simple and solvable are the watchwords. There is no advantage in representing a complex system with a complex model, nor is there in replacing one unsolvable system with another. An important requirement for a model is that it be self-consistent.

It took a model like SIR to identify some of the important thresholds in an epidemic evident (22, 23). Weiss (24) explains why both the threshold for infection and the herd immunity threshold are difficult to deduce from data.

Simple though the models are, the truth and beauty in them are compelling and emerge only from exact solutions. It is prudent to extract as much information from models as possible before trying to modify the model and introduce new unknown parameters that can quickly turn it into a curve fitting experiment that does not illuminate the process.

The number of unknown parameters is inversely proportional to the complexity of a model and so a desirable feature would be that the models have the least number of unknown parameters. If data do not match the results of a model, and that happens often, then it is an inappropriate use of the model, and it is time to move on to a new model. The model is not “wrong.”

### Numerical solutions can be a trap

But models are not a panacea either. In the case of the Covid 19 SIR model, the equations of the model are nonlinear and the solutions are approximate. Hirsch et al. (11) and Strogatz (12) point out the familiar mathematical traps related to numerical solutions to nonlinear equations. The Universal rule discovered here, for example, would most likely remain hidden, if only numerical solutions were available. Exact solutions in the form of analytic functions makes analysis possible, and that helped reveal the Universal rule.

### Important prior work

After this work was complete, we became aware of two other papers that produced analytic solutions earlier. Harko et al. in 2014 performed multiple transformations and magically deduced the analytical result, but in the process, the relationship to the true time variable became complicated. Miller in 2012, also derives (26) the analytic solution in a more direct fashion, but his interest seems to be in connecting SIR to edge-based compartmental models. For reasons that are not clear, these insightful papers of Miller and Harko et al. have not received the attention they merit, and further, no one seems to have advanced their analyses.

### Why is there a Universal rule?

A heuristic explanation for the Universal rule would be to observe that this population is a “closed” system, where the only two events that happen are infection and removal and one affects the other. We would expect them to be somehow related, not necessarily in a linear fashion, of course.

A better but more technical explanation is as follows. The model parameters are algebraically related in an ideal situation. But exogenous factors, individual to each of the countries and states, mask the relationship by introducing a time dependence in them. There is, however, a holonomic constraint (27) evident, for every time-step, in the form of the universal linear rule that relates the two parameters of the model to observables.

*It is worth noting that* Changes in both 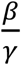 and 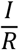 remain correlated but they are not monotonic. As social distancing and mask wearing get enforced, both I and R change, and to see it as a time series in Tableau (28) is mesmerizing. It is possible to use Tableau to correlate specific events, such as shutdowns in a state, with the changes in both 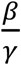 and 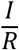.

### Complexity or simplicity

As stated earlier, the initial attempts at finding an empirical rule were by trial and error. For example, 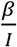 and 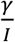 when plotted on X and Y axis respectively, were linear in some cases and piecewise linear in others. What if the choice made first were 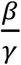 and 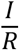 leading to the accidental discovery of the Universal rule? That would have been akin to the discovery of the Gutenberg Richter law (4) describing the frequency of earthquakes of different magnitudes, or any of the other patterns seen in studies of complex systems (3 - 6). There would then be claims of self-organized criticality in the pandemic, and the universal rule embraced as another example of complexity in our lives. But that’s not what happened.

The success in fraud detection through linearization encouraged a different path for the analysis. Deriving the exact analytic solution of a deterministic dynamical system, combined with an adaptation of Emmy Noether’s theorem, led to the universal rule. So, which is it? Is the universal rule of the Covid 19 pandemic an example of a self-organized criticality of a nonequilibrium system or just another example of truth and beauty in a simple deterministic model? It’s anyone’s guess.

### Universal rule should apply to other epidemics and models

The universal rule is a consequence of the SIR model, and so, other short-lived epidemics where the SIR model was used with success, ought to exhibit the same linear law. This ought to be verified but is beyond the scope of the paper. That the Covid 19 global data comply with the universal rule only means that the SIR model is a reasonable approximate mathematical model to describe the pandemic. But there is room for improvement.

### Limitations of the model

The limitations of the SIR model are well documented and many of them inspired the formulation of other, more advanced and complex compartmental models. As pointed out earlier, adding more and more unknown parameters is not always prudent. Often the causal connections between the parameters are lost and then it becomes sheer curve fitting, not science. Of course, exogenous factors are out of control but there are specific features, like modeling the change in transmission rates, when more recovered people mix with the susceptible population could prove useful. These will challenge homogeneous mixing assumptions and perhaps help clarify the time dependence introduced, particularly in *β*. Clearly, for good forecasts there is a need for new and improved epidemic models that captures the time dependence of parameters, even while complying with the Universal rule.

#### Stopping the pandemic - Different options

There are three distinct ways to stop the epidemic.

i. *Lower infections to control the epidemic:* Social distancing, mask wearing, quarantining and contact tracing are all known methods to reduce infection. Many states took these suggestions seriously while others did not. Absent quantitative measurements it is difficult to argue for or against them. Coincidentally, Atul Gawande in an article in the New Yorker (29) called North Dakota the “worst hit state in the worst hit country in the world”. Our analysis shows that North Dakota is also the best state in curbing infections and reversing the epidemic in the nation. It is difficult to make any generalizations, since the states that have done well in keeping infected populations low are those that are not fans of CDC guidelines or mask mandates. We have absolutely no explanation as to why, for example, South Dakota, that hosted a 500,000 strong motor cycle rally at Sturgis, and openly flouted best practices of mask wearing and social distancing, found its infected population decreasing substantially and reversing the epidemic by January 2021. It seems as if Covid 19 defies simple categorization.
ii. *Promote runaway infections, as in Sweden:* As we found, that was not successful and the entire program was considered a failure at many levels. But as before, Covid 19 fights stereotyping. A counter example of sorts, is the case of the cruise ship Diamond Princess, with 2670 total passengers and 712 in the Removed category, they reached Herd immunity with 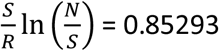. This was a case of runaway infections leading to Herd immunity – as Sweden hoped. So, it seems to work, but not in all cases. Speculating for a moment, it could be that in the narrow confines of a cruise ship, the rate of spread was very fast and very quickly the condition 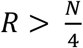 (that is used for vaccination programs) was met and so it was not subject to the condition *S* < *S*_2_ (with 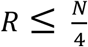) to reach Herd immunity.
iii. *Herd immunity through vaccinations:* The Universal rule imposes a mathematical restriction that Herd immunity begins around the time when 25% of the total population has been vaccinated. This is a mathematical conclusion that is startlingly different from the current consensus that 70% and more must be vaccinated to trigger Herd immunity. This also means that President Biden’s promise of 100 million people vaccinated in the first 100 days is actually a path to imminent Herd immunity in many states of the nation. It is not as if vaccination is the only way to reach the onset condition. As long as 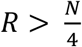, all values of S satisfy the inequality to reach Herd immunity, as discussed in the case of the Diamon Princess cruise ship. The situation in Dharavi, the crowded slum in Mumbai, is perhaps another example of runaway infections building Herd immunity. In Dharavi, Covid 19 cases have plummeted without vaccination. One explanation offered is that it reached Herd immunity via infections, and the population being younger, the community escaped with minimal collateral damage of hospitalizations and death. The measured seroprevalence in Dharavi was 57% which is quite higher than our 25% minimum, and we would also conclude that Dharavi reached Herd immunity. But why did Dharavi reach such a limit while Sweden struggled to go higher than 7%? Is it because Dharavi, with its super crowded community, accelerated the spread of the disease — not by choice, of course — and reached the 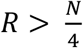 while Sweden adopted a passive route and was stuck at a low level far below Herd immunity levels?

### Will onset of Herd immunity lead to the end of the pandemic?

Probably not. Even though the pandemic is abating, it has not been eradicated. The residual infected populations can continue to infect, albeit at a lower rate. Continued mass vaccinations can off-set this reemergence of the epidemic and restore Herd immunity.

An interesting way of looking at this is as follows. If we define “confirmed,” as (infected+ removed), the ratio of “infected” to “confirmed” is a useful metric for gauging potential residual damage to a population after onset of Herd immunity. In the South East Asian countries that practiced mask wearing and quarantining, this ratio currently tends to be 30% or lower. Assuming the Johns Hopkins numbers are correct, the same ratio in about 19 US states is higher than 80%, with half including IL, KS, NV, FL, GA and MO even higher than 97%. In other words, these US states have managed to lower the daily new cases, but there is a large pool of infected that is still present, and the virus is slowly working through the population. Perhaps it is useful to define a “half life” for the residual infected populations, as a way to rate the efficiency of states in controlling further infections.

The onset point 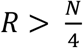 is then to be interpreted as a possible start point of the decay of the epidemic – but not as a confirmation of wiping out the disease. Maintaining Herd immunity is critical to prevent super-spreader events that revive the epidemic.

#### Summary and conclusions

The rescaled SIR model equations led to an exact analytical solution which in turn revealed a new Universal rule for Covid 19 data. This met the gold standard for a model because it predicted an unknown and new rule that was later confirmed through observations.

There is an opportunity during the early stages of the epidemic to control the growth of the epidemic by lowering the Infected population, as North Dakota and several other states have demonstrated. As to why these communities succeeded even while not following CDC guidelines is a mystery.

As more get infected, the Universal rule reveals a negative feedback effect against runaway infections, slowing down the spread, making it mathematically improbable to develop herd immunity through mass infections, as Sweden learned recently. But the inadvertent success on the Diamond Princess and Dharavi makes us pause before dismissing the runaway infections as a feasible path to Herd immunity.

Most intriguing, the Universal rule makes a new prediction that vaccination of just about 25% of the total population can trigger onset of Herd immunity. If President Biden’s promise of 100 million vaccinations in the first 100 days holds true, then this prediction will also be tested soon.

But maintaining the immunity, going forward, will require a serious effort from the states because at this point there is still a large pool of infected patients in many states, and there is a temporary stasis, a balance between infections and removals. If the normal practices of mask wearing and social distancing are not continued for another few months, there can be super-spreader events that revive the epidemic, requiring many more to be vaccinated. And the emergence of variants and new strains of the virus can further complicate matters.

## Supporting information

global covid data for tables 1 and 2

## Data Availability

All the necessary data to reproduce the results are in an excel file that is in the supplemental files.

## Acknowledgments

I wish to thank Professor N. Mukunda for an erudite explanation of holonomic constraints in classical mechanics, Dr. Nathaniel Cobb for the numerous discussions on chronic diseases and epidemics and Professor Howard Weiss for his comments on Herd immunity. I also wish to thank P. K. Rajashekhar for extracting the data from Johns Hopkins, calculating the various integrals using MATLAB and assisting in creating the many graphs supporting the Universal rule. This work would not have been possible without the relentless effort of Professor Lauren Gardner and her student, Ensheng “Frank” Dong of the Center for Systems Science and Engineering at the Johns Hopkins University, in making the most current global Covid 19 data available at no cost to the public. It is my fervent hope that Johns Hopkins University and other organizations like the WHO, are funded continuously and generously, so as to prevent commercial organizations from gaining exclusive access to the pandemic data and monetize our misery. Lastly, I wish to dedicate this paper to the memory of my old friend and colleague, Professor Sandip Pakvasa, former Chairman, Department of Physics, University of Hawaii, who passed away on September 24, 2020.

## Notes

### Competing Interest Statement

The authors have declared no competing interest.

### Funding Statement

There was no external funding for this work.

### Author Declarations

There were no relevant ethical guidelines as the research dealt only with public and demographic global data as published by Johns Hopkins

